# RNA sequencing identifies a cryptic exon caused by a deep intronic variant in *NDUFB10* resulting in isolated Complex I deficiency

**DOI:** 10.1101/2020.05.21.20104265

**Authors:** Guy Helman, Alison G. Compton, Daniella H. Hock, Marzena Walkiewicz, Gemma R. Brett MGenCouns, Lynn Pais, Tiong Y. Tan, Ricardo De Paoli-Iseppi, Michael B. Clark, John Christodoulou, Susan M. White, David R. Thorburn, David A. Stroud, Zornitza Stark, Cas Simons

**Affiliations:** Murdoch Children’s Research Institute, Royal Children’s Hospital, Victoria, Australia; Institute for Molecular Bioscience, The University of Queensland, Queensland, Australia; Department of Paediatrics, University of Melbourne, Melbourne, Victoria, Australia; Department of Biochemistry and Molecular Biology and Bio21 Molecular Science and Biotechnology Institute, University of Melbourne, Victoria, Australia; Victorian Clinical Genetics Services, Murdoch Children’s Research Institute, Royal Children’s Hospital, Victoria, Australia; Center for Mendelian Genomics, Broad Institute of MIT and Harvard, Cambridge, Massachusetts, USA; Centre for Stem Cell Systems, Department of Anatomy and Neuroscience, The University of Melbourne, Victoria, Australia

## Abstract

The diagnosis of mitochondrial disorders remains a challenging and often unmet need. We sought to investigate a sibling pair with suspected mitochondrial disease and a clinical presentation notable for global developmental delay, poor growth, sensorineural hearing loss, and brain MRI abnormalities, both with early death. Following uninformative exome and genome sequencing of the family quartet, RNA sequencing was pursued as an orthogonal testing strategy. RNA sequencing of fibroblasts from the older sibling identified the presence of a cryptic exon in intron 1 of *NDUFB10*, that included an in-frame stop codon. *NDUFB10* encodes a subunit of mitochondrial OXPHOS complex I. Differential expression analysis relative to control samples suggested significantly decreased expression. The cryptic exon was found to contain a rare intronic variant, NM_004548.3:c.131-442G>C, that was homozygous in both affected siblings and absent from population allele frequency databases. Immunoblot and quantitative proteomic analysis of fibroblasts from the older sibling revealed decreased abundance of complex I subunits associated with NDUFB10, providing evidence of isolated complex I deficiency. Biallelic variants in *NDUFB10* have previously been reported in a single individual with infantile-onset mitochondrial disease. We present data implicating a deep intronic variant in *NDUFB10* as the cause of mitochondrial disease in two further individuals. This variant results in loss of expression and overall destabilization of mitochondrial OXPHOS complex I and highlights the importance of RNA sequencing as a complementary diagnostic tool in patients undergoing genome-wide diagnostic evaluation.

## Main Text

Mitochondrial diseases are genetic disorders with a primary defect in oxidative phosphorylation (OXPHOS), an important cellular energy generating system [5,9]. The majority of mitochondrial disease-associated genes are related to OXPHOS biogenesis (185 of 289 genes) [5]. OXPHOS requires the assembly of five unique enzyme complexes, complex I – complex V, with complex I (NADH:ubiquinone reductase) being the largest of these with 44 unique subunits and more than 15 assembly factors [18], 27 of which have been associated with mitochondrial disease [5, 15]. Most subunits are nuclear-encoded, however seven subunits are encoded on the mitochondrial genome. While the rate of gene discovery has accelerated substantially in the genomic era, diagnosis can still remain challenging in some cases. For example, pathogenic DNA variants in non-coding regions can often be difficult to identify using standard genome analysis approaches.

Variants in *NDUFB10*, a complex I subunit, have previously been reported in only one individual, an infant with fatal lactic acidosis and cardiomyopathy and decreased complex I activity in multiple tissues including muscle, heart, and liver [6]. Exome sequencing identified compound heterozygous variants in *NDUFB10*, a nonsense variant *in trans* with a missense variant. To our knowledge, this is the only report of mitochondrial disease related to dysfunction of *NDUFB10*. We describe a sibling pair with a clinical presentation suggestive of mitochondrial disease with hearing loss, neurological deterioration, and early death. Exome and genome sequencing of both affected individuals was unable to identify a molecular diagnosis. Using RNA sequencing as part of tiered-diagnostic analysis, we detected aberrant splicing in an intronic region of *NDUFB10*, with markedly decreased expression of this gene relative to controls. We present transcriptomic and proteomic data implicating a biallelic deep intronic variant as the cause of complex I deficiency in these individuals.

All research activities were performed under institutional ethics approval from The Royal Children’s Hospital Melbourne (HREC/16/RCHM/150) and written informed consent was provided by both parents. Individuals 1 and 2 are siblings, born of fourth-degree consanguineous parents of Pakistani origin (Table 1). There is one unaffected sibling with a history of ventricular septal defect noted at birth, but otherwise no noted family history of neurologic disease. The mother had a history of hypothyroidism and there are two paternal cousins with hearing impairment.

**Table 1.**
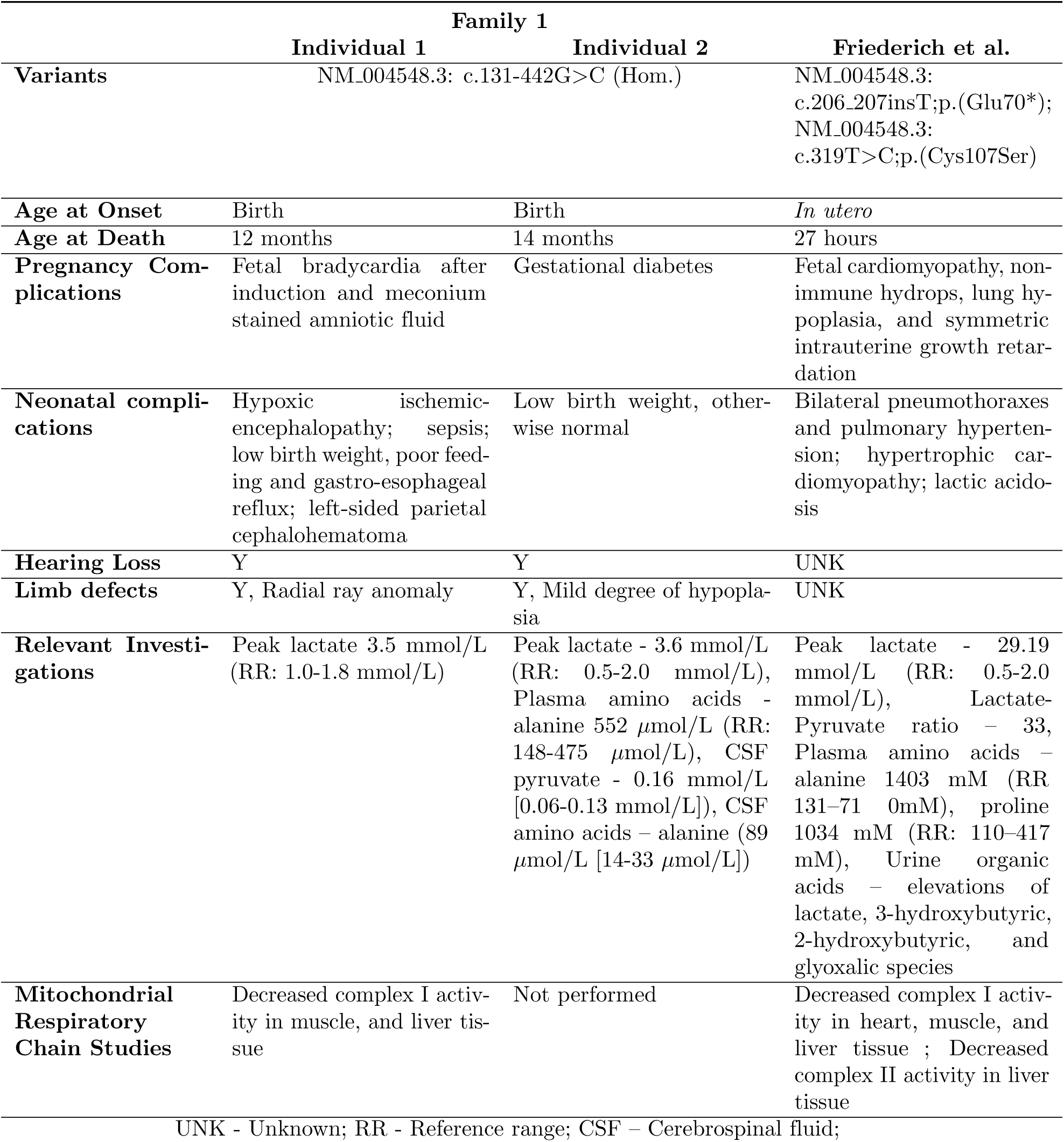
Clinical characteristics of *NDUFB10*-affected individuals.

Individual 1 is the elder of the affected sibling pair. He presented at birth, with delivery by emergency Cesarean section due to bradycardia after induction and meconium stained amniotic fluid. He stayed in the neonatal intensive care unit for 26 days due to hypoxic ischemic-encephalopathy, low birth weight (2.72kg [<10th percentile]) and sepsis, in addition to poor feeding and gastro-esophageal reflux. MRI revealed a left-sided parietal cephalohematoma due to birth trauma. A skeletal survey revealed shortened first metacarpals bilaterally and gracile osteopenic parietal bones. At five months of age, he was noted to have slow growth, with a weight of 4.74 kg (<3rd percentile) and a head circumference of 40 cm (<3rd percentile). Auditory brainstem response testing diagnosed absent right-sided hearing and profound high frequency hearing loss on the left at seven weeks of age. In the months after discharge he had developed good head control and smiled at three months. On examination he was bright and alert. He had simple low-set ears and bilateral single palmar creases. He had a flexion deformity of the left hand (Type 1 radial dysplasia) and hypoplasia (Type II) of the left thumb, suggestive of a radial ray anomaly (Figure 1:1C). At nine months of age, he was still below the 3rd centile in all growth parameters and had made some development gains in gross and fine motor skills. He made babbling noises and had become attentive to voices. He had multiple admissions due to concerns with growth and feeding, abnormal posturing episodes, and difficulties with respiration. At 11 months of age he was admitted due to increased episodes of apnea requiring intubation and died from complications of his underlying disease.

**Figure 1.**
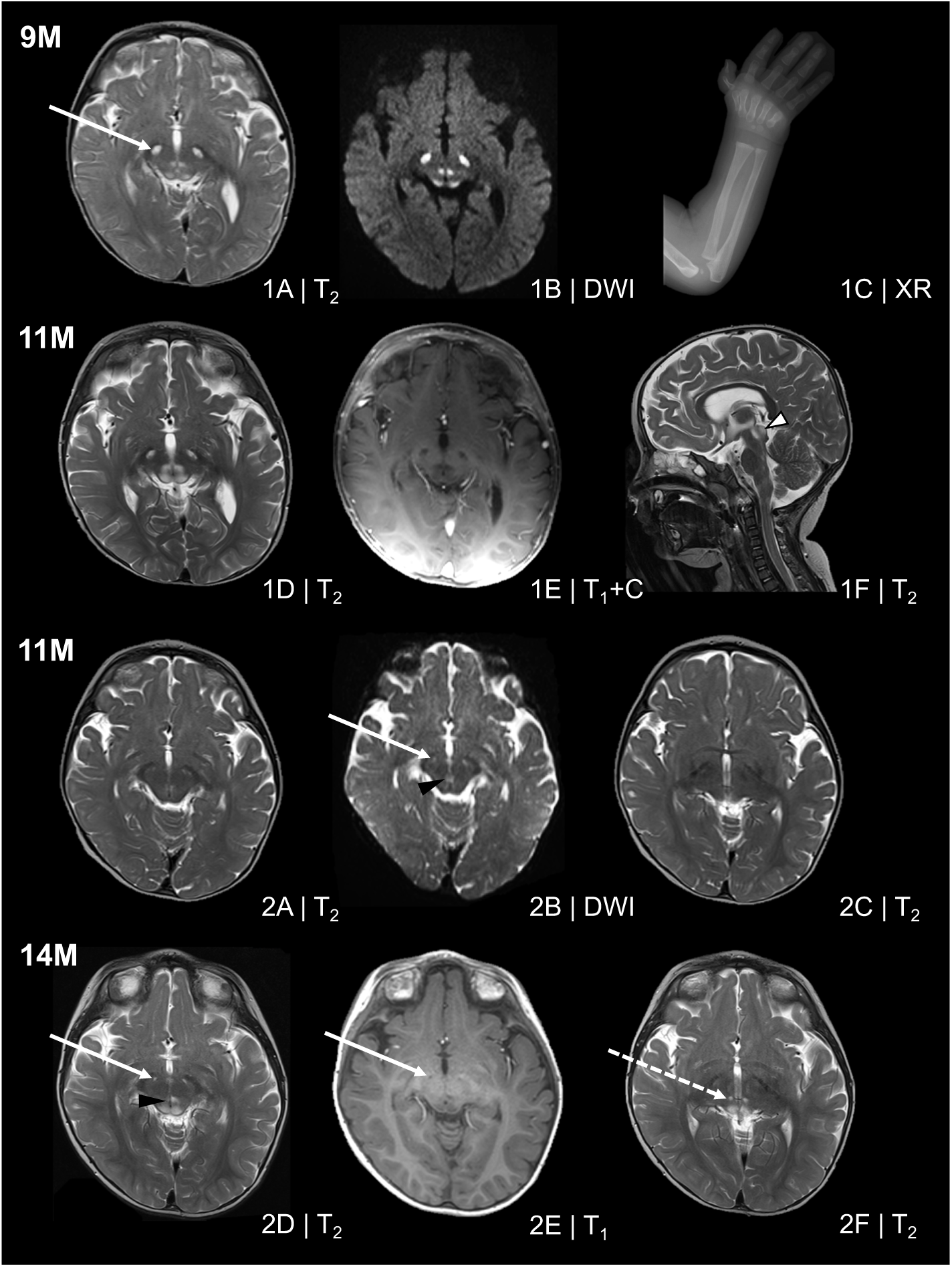
Imaging of *NDUFB10* affected individuals. Magnetic resonance imaging was performed according to standard clinical protocols at nine and eleven months in Individual 1 and eleven and fourteen months in Individual 2. (1A-1B) Imaging at nine months of age in Individual 1 revealed T_2_ hyperintensity in the cerebral peduncles predominantly affecting the corticonuclear (white arrow) and frontopontine tracts, the peri-aqueductal midbrain and the inferior colliculi. Hypointensity on T_1_ (not shown) and bright regions on diffusion weighted imaging suggested a demyelinating process. There was no restricted diffusion confirmed on the apparent diffusion coefficient (ADC) maps (not shown). Myelination was appropriate for age. There was mild enlargement of the ventricles and extra-axial spaces suggestive of atrophy. (1C) A left forearm X-ray performed at the same age shows hypoplasia of thumb and a mild flexion deformity. (1D-1E) Progression of the neuroimaging features observed in Individual 1 was observed on follow-up imaging at 11 months of age. There is continued hyperintensity on T_2_, and diffusion weighted imaging and hypointensity on T_1_ with (1E) and without contrast. There was extension superiorly to the superior colliculi (white arrowhead with black outline, 1F) and inferiorly into the dorsal pons and medulla. (2A-2C) Neuroimaging of the similarly affected sibling reveals similar features albeit less significant involvement at matched time points. (2B) The corticonuclear fibres (white arrow) and periaqueductal midbrain (black arrowhead) are mildly affected although not to the degree of the similarly affected sibling. Follow-up imaging shows a progression of these features, with extension into the inferior colliculi of the midbrain. (2E) A demyelinating process was suspected based on T_1_ images showing hypointensity within the corticonuclear fibres, peri-aqueductal midbrain, and the inferior colliculi. There are isolated lesions of the inferior thalami (white dashed arrow, 2F) and superior colliculi that were not apparent on imaging three months prior (2C)

Laboratory investigations including urine organic acids and glycosaminoglycans, plasma very long chain fatty acids, amino acids and serum transferrin isoforms, were all normal. Plasma lactate levels were elevated at 7 months (3.5 mmol/L [1.0-1.8 mmol/L). Pyruvate dehydrogenase complex activity in fibroblasts was normal. Respiratory chain enzyme activities were normal in fibroblasts. In skeletal muscle, when enzymes were expressed relative to protein, Complex I activity was normal but Complex II, III, and IV activities were elevated. Hence, Complex I activity was borderline low (40% residual activity) relative to citrate synthase and Complex II. Mitochondrial complex I activity was also borderline low in liver relative to citrate synthase (50% residual activity while Complex II, III, and IV activities were in the range 87 to 120%). A muscle biopsy showed no significant histological pathology. Electron microscopy from a liver biopsy was unremarkable.

Further MRI studies were performed at nine months in Individual 1. There were signal abnormalities in the brainstem with T_2_ hyperintensity and T_1_ hypointensity suggesting demyelination in the corticonuclear tracts within the cerebral peduncles and affecting the inferior colliculi (Figure 1:1A-1B). The peri-aqueductal midbrain was affected. There was prominence of the ventricles and extra-axial spaces. On repeat studies two months later, the signal abnormalities in the midbrain were more extensive, affecting the substantia nigra and locus ceruleus as well as the superior colliculi (Figure 1:1F) and extending inferiorly into the dorsal pons and medulla. MRS did not reveal elevated lactate in the basal ganglia but technical limitations precluded this from being performed in the brain stem.

Individual 2 is Individual 1’s younger brother who was born at term by repeat elective Cesarean section. The pregnancy was complicated by gestational diabetes mellitus requiring insulin in the final weeks of the pregnancy and maternal vitamin D and iron deficiencies and hypothyroidism requiring thyroxine. His birth weight was 2.9 kg (25th percentile) and head circumference was 33 cm (<10th percentile). His newborn screening hearing test and perinatal period was normal. He made developmental gains without regression and at seven months was able to sit independently, roll over, transfer objects, vocalize, and was attentive to objects and sounds. At seven months he was noted to have a unilateral auditory neuropathy on audiologic testing which progressed bilaterally by eight months. Independent sitting was noted to have been lost on follow-up at nine months of age. On examination, he had failure to thrive and laryngomalacia with an inspiratory stridor. Extraocular eye movements were intact. There was mild head lag and truncal hypotonia with increased peripheral tone. He had good strength but reflexes were depressed. There was no clonus. At 14 months of age he was admitted to the intensive care unit due to episodes of bradycardia and persistent apnea, and noted to have increasing dystonic movements. He died at 14 months of age.

Laboratory studies included elevated blood lactate (3.6 mmol/L), normal creatine kinase and normal thyroid stimulating hormone. Plasma amino acids showed a mildly elevated alanine at 552 *μ*mol/L (Reference range:148-475 *μ*mol/L). Urine organic acids showed persistent excretion of fumaric acid. Very long chain fatty acids and serum transferrin isoforms were normal. Cerebrospinal fluid (CSF) pyruvate was mildly elevated (0.16 mmol/L [0.06-0.13 mmol/L]) and CSF alanine was significantly elevated (89 *μ*mol/L [14-33*μ*mol/L]). No mitochondrial respiratory chain studies were performed on this individual.

An MRI at 11 months revealed T_2_ and FLAIR signal changes symmetrically within the cerebral peduncles, peri-aqueductal midbrain (Figure 1:2A-2B), and also at the anterolateral aspect of the 4th ventricle. On follow-up studies at 14 months, there was progression of the midbrain findings (Figure 1:2D-2E). There was new involvement of the inferior and superior colliculi and symmetrical small round areas of T_2_ hyperintensity within both medial thalami inferiorly (Figure 1:2F). These findings were consistent, albeit less severe than his similarly affected sibling.

A chromosomal microarray was negative and breakage analysis to assess for Fanconi Anemia performed in the elder sibling was normal. Single gene analysis of *GJB2* for a genetic cause of the progressive hearing loss observed in Individual 1 was also normal. Testing for common mitochondrial genome point mutations and exome sequencing of both affected siblings and biological parents did not identify any candidates felt to be relevant to these patient’s phenotype upon multidisciplinary review. Genome sequencing was then performed on both affected siblings and their parents by the Broad Center for Mendelian Genomics but was also unable to identify variants in phenotype-compatible genes.

Human whole transcriptome sequencing on RNA derived from fibroblasts of Individual 1 was performed by the Genomics Platform at the Broad Institute of MIT and Harvard. The transcriptome product combines poly(A)-selection of mRNA transcripts with a strand-specific cDNA library preparation. Libraries were sequenced on the HiSeq 2500 platform to generate 50 million 2 × 100 nt reads. Following alignment to the Human Reference Genome Build 38 using STAR (Version 2.7.3a, [4]), duplicate reads were masked with Picard MarkDuplicates and quantification was performed using FeatureCounts from the R Subread package (Version 1.34.7, [13]). Gene level differential expression analysis was performed using the DESeq2 package (Version 1.25.9, [14]).

Differential expression analysis identified a greater than three-fold reduction in expression of the nuclear encoded mitochondrial gene *NDUFB10* relative to 24 similarly sequenced controls (Log_2_ Fold Change -1.81; p-adj = 9.058787e-06 [Figure 2A; Supplemental Table S5]). Inspection of the reads mapping to *NDUFB10* identified two novel splicing events consistent with the inclusion of a 94-nt cryptic exon (referred to as exon 1A) within intron 1 of *NDUFB10*. Neither of the novel junctions were observed in any of the 24 control samples and no transcripts utilizing either junction are present in either the Gencode v32 or Refseq human transcript databases. Approximately 32% of junction-spanning reads using the exon 2 acceptor splice site support the inclusion of the cryptic exon in Individual 1. In addition to the cryptic exon, reads mapping along the length of intron 1 were enriched in Individual 1 relative to controls, suggesting that intron 1 may be retained in some transcripts. The inclusion of exon 1A would result in a frameshift p.(Arg43fs^*^32), while intron retention would be predicted to introduce a premature termination sequence p.(Arg43fs^*^135) to this transcript, with both events expected to be subjected to nonsense-mediated decay (NMD) leading to an overall reduced level of *NDUFB10* in this individual.

**Figure 2.**
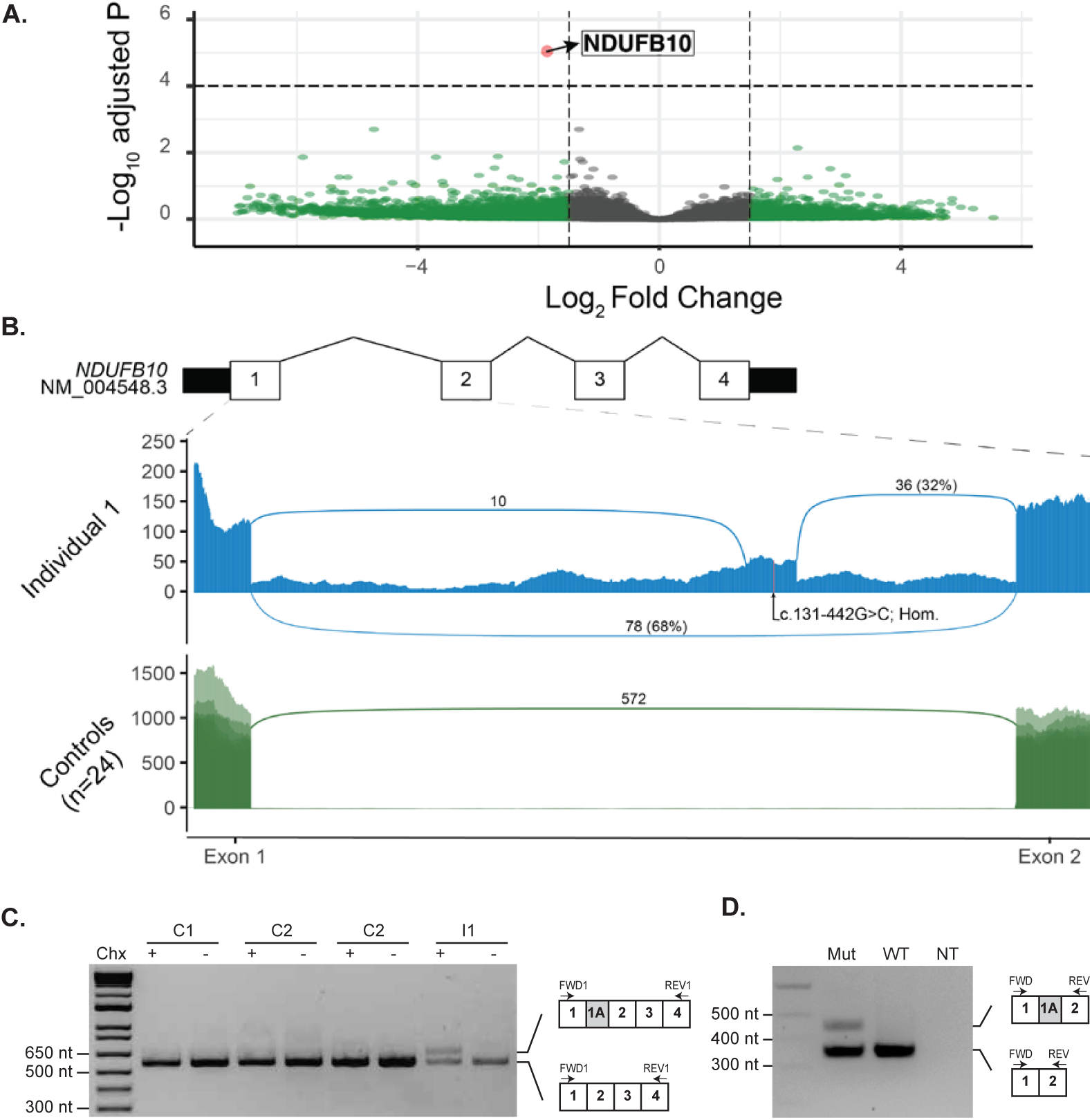
Identification of reduced expression and aberrant splicing in Intron 1 of NDUFB10 in RNA sequencing data from Individual 1. (A) Analysis of RNA sequencing data revealed significantly decreased expression of *NDUFB10* in the affected individual compared to 24 similarly sequenced controls. Dashed lines represent a Log_2_ Fold Change (L_2_FC) cut-off of 1.5 or -1.5 on the x-axis and by the log transformed adjusted p-value on the y axis. Details of each gene meeting these thresholds are provided in Supplemental Table S5. *NDUFB10* is labeled in red and can be seen as a clear outlier with a L_2_FC of -1.81 and p-adj = 9.06e-06. Genes colored in green were significant based on their L_2_FC values as described above while genes color in grey were not significant for their adjusted p-value or for the L_2_FC value. (B) Sashimi plot depicting junction spanning reads and read depth at each nucleotide from exon 1 to exon 2 of *NDUFB10*. RNA sequencing data from fibroblasts of Individual 1 are shown in the top panel. Two novel splice junctions can be seen suggesting the presence of a cryptic exon in intron 1. The position of the homozygous variant, c.131-442G>C is shown near the center of the putative cryptic exon. The lower panel displays an overlay of 24 similarly sequenced fibroblast control samples. No reads supporting either of the novel splice junctions were detected in any of the control samples. (C) RT-PCR of *NDUFB10* from fibroblast RNA from Individual 1 (I1) and three control lines (C1, C2 and C3) with (+) or without (-) treatment with cycloheximide (Chx). (D) A mini-gene splicing assay containing the c.131-442G> C was prepared and amplification of RNA from both the mutant and wild-type plasmids by RT-PCR using primers between exon 1 and exon 2 of *NDUFB10* yielded an amplicon consistent with the size expected from reference *NDUFB10* splicing. In the mutant plasmid, a second larger amplicon was present. This amplicon, approximately 94-nt in size, is absent in the wild-type plasmid and is consistent in size with aberrant splicing event observed on RNA sequencing data.

To confirm the presence of a cryptic exon, we used reverse transcription polymerase chain reaction (RT-PCR) to amplify transcripts from fibroblast RNA using primers situated in exon 1 and exon 4 of *NDUFB10* (Supplemental Table S1). An amplicon consistent with the expected size of 563-nt was generated from Individual 1 as well as fibroblasts from three healthy controls (Figure 2C). An additional larger amplicon consistent with the inclusion of the cryptic exon observed in the RNA sequencing data was amplified from Individual 1, but only when fibroblasts were treated with cycloheximide to inhibit NMD, consistent with the hypothesis that this transcript is subject to degradation. The larger amplicon was not detectable in any of the three control lines.

To confirm the sequence and genomic location of the cryptic exon observed by RNA sequencing we used an Oxford Nanopore Flongle to sequence full length reads of the RT-PCR amplicons from cycloheximide treated fibroblast RNA Over 5,000 reads were generated from Individual 1 and three healthy controls. The reads were processed using the Full-Length Alternative Isoform analysis of RNA (FLAIR; [19]) analysis tool to identify and quantitate the isoforms present in each RT-PCR. An isoform containing the cryptic exon identified by RNA sequencing was present in 24.5% of reads from Individual 1 (Supplemental Figure S1). No nanopore reads were identified that supported the retention of intron 1 that was observed in the RNA sequencing data from Individual 1. However, amplification of transcripts retaining intron 1 maybe disfavored by RT-PCR due to their increased length which could explain their absence from the sequencing library.

With these data in mind, we reviewed the genome sequencing data to identify variants consistent with recessive inheritance in *NDUFB10*. In intron 1 there were 17 variants that were homozygous in both affected individuals; of these 16 could be excluded on the basis of high population allele frequency (>0.05) and previously observed homozygotes seen in a reference allele database (Supplemental Table S6). The only remaining variant, NM_004548.3:c.131-442G>C, sits within the cryptic exon (Figure 2B) and was found to be homozygous in both affected siblings and heterozygous in the unaffected parents. This variant is absent in available population allele frequency databases and internal datasets and is conserved across primate species.

To determine if the c.131-442G>C variant was responsible for the observed *NDUFB10* splicing defect we generated a mini-gene splicing reporter by cloning 2.1 kb of genomic sequence, including exon 1 to exon 3 of *NDUFB10*, into the mammalian expression vector pEYFP-C1. Primer directed mutagenesis was used to generate a version of the plasmid containing the c.131-442G>C variant before transfection into human embryonic kidney (HEK293T) cells. Amplification of RNA transcribed from both the mutant and wild-type plasmids by RT-PCR using primers between exon 1 and exon 2 of *NDUFB10* yielded an amplicon consistent with the size expected from reference *NDUFB10* splicing (Figure 2D). A second approximately 100-nt larger amplicon was generated only by the mutant plasmid, consistent with the aberrant splicing observed by RNA sequencing in Individual 1.

We sought to determine if the aberrant *NDUFB10* splicing affects the overall stability of complex I or other mitochondrial respiratory chain complexes. Immunoblotting was performed using an antibody against one subunit of each of the five mitochondrial OXPHOS complexes on cell lysates isolated from a fibroblast sample from Individual 1, an affected patient control with known complex I deficiency due to a homozygous variant in NDUFB3 (OMIM:603839) [1] and three control samples. This revealed an absence of the complex I subunit NDUFB8 and normal levels of the remaining mitochondrial OXPHOS complexes relative to controls (Figure 3A), consistent with an isolated complex I defect.

**Figure 3.**
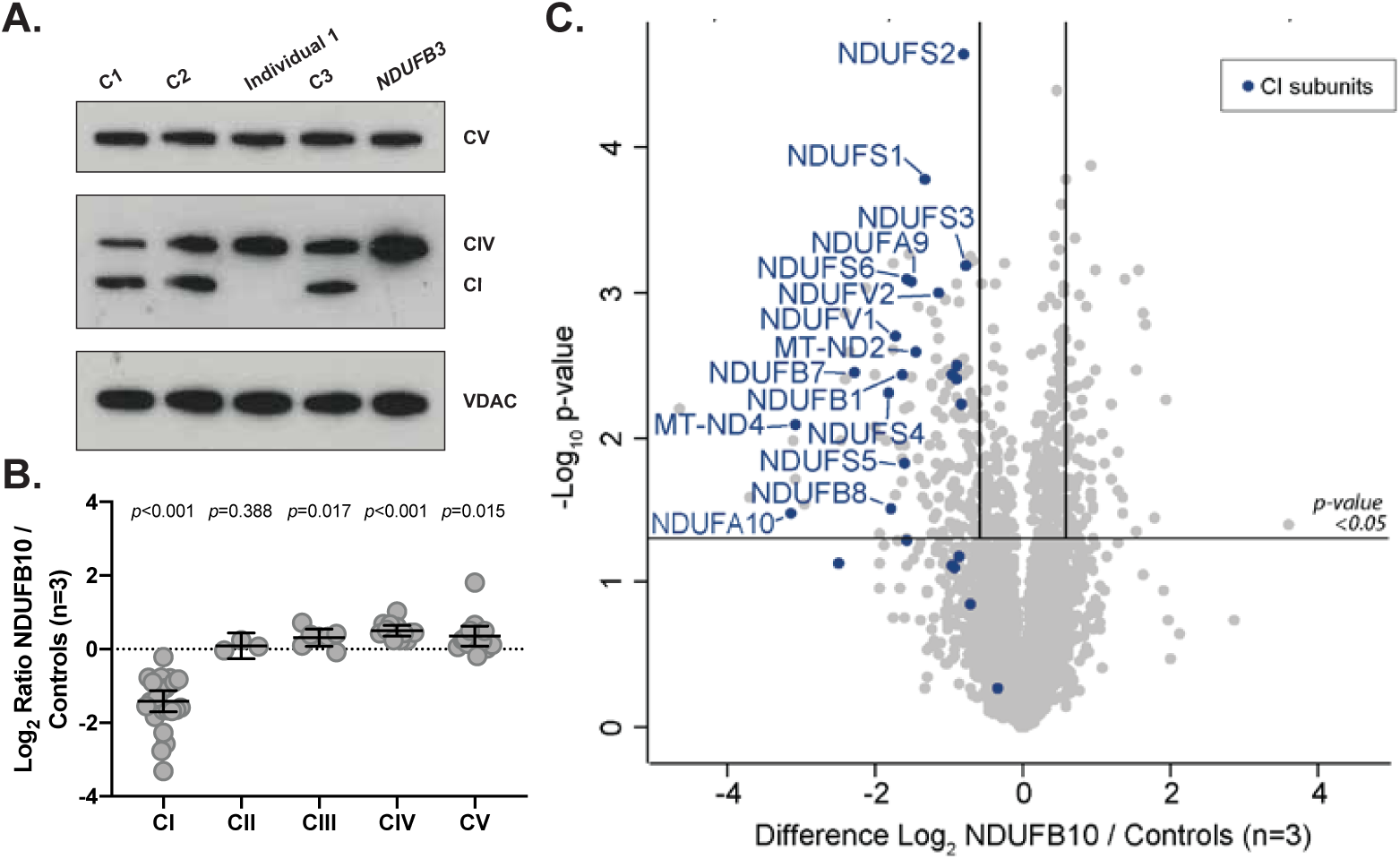
Evidence of isolated complex I deficiency in Individual 1. (A) SDS-PAGE and immunoblotting of protein isolated from fibroblasts and probed with antibodies specific for mitochondrial OXPHOS proteins reveals an absence of complex I subunit NDUFB8 in Individual 1 from the family in this study (Lane 3) relative to controls (Lane 1, 2, 4) similar to a patient with known complex I defect due to variants in NDUFB3 previously reported in Calvo et al. [1] Levels of COX2 (complex IV; CIV) and ATP5A (complex V; CV) and VDAC1 as the loading control are also shown. (B) Analysis of OXPHOS protein levels detected using quantitative proteomics in Individual 1, represented as Log2 ratio of the mean of NDUFB10 and controls. The middle bar represents the mean value, while the upper and lower bars represent the 95% confidence interval of the mean value. Each dot represents a single protein. (C) Volcano plot depicting quantitative proteomics data revealing decreased levels of complex I subunits in Individual 1 compared to controls. The horizontal line within the volcano plot represents a significance value of p=0.05 whereas the vertical lines represent fold-change of +/-1.5. Complex I subunits are depicted in blue.

Having identified an isolated complex I defect due to transcription of a cryptic exon in *NDUFB10*, quantitative proteomic analysis was then performed on fibroblasts from Individual 1 relative to three individual controls to determine the effects of the mutation on the cellular proteome. We detected over 5,000 unique proteins and quantified the changes in 4,200. While NDUFB10 was readily detected in all three of the controls, it was only detected in one of the three replicates from the patient and with a greater than 4 fold reduction in abundance (Supplemental Table S7 and S8). Moreover, the abundance of the majority of complex I subunits were decreased in Individual 1 relative to controls while the levels of other mitochondrial respiratory chain complex subunits remained relatively unchanged (Figure 3B and C). These results are largely consistent with our previously reported study of a HEK293T cell line gene-edited to lack expression of NDUFB10 [18], suggesting the effect of the variant in Individual 1 leads to similar defects in assembly of the complex (Supplemental Figure S2).

In this study, we used RNA sequencing to identify an underlying etiology of disease in an extensively investigated family with previously inconclusive exome and genome sequencing. RNA sequencing identified aberrant splicing leading to significantly reduced expression, guiding reanalysis of genome sequencing to a deep intronic variant in *NDUFB10*. Through SDS-PAGE and immunoblotting and subsequently quantitative proteomics, we were able to identify that the molecular defect caused by this variant resulted in destabilization of complex I in a similar way to that observed in gene-edited HEK293T cells lacking NDUFB10. Importantly, HEK293T cells lacking NDUFB10 also exhibited no detectable complex I enzyme activity although they had severe defects in mitochondrial respiration [18]. The data provided here further support evidence that genetic variants in *NDUFB10* result in defective assembly of complex I and clinically manifest as severe mitochondrial disease.

To our knowledge, only a single case of mitochondrial disease associated with variants in *NDUFB10* has previously been reported. Friederich et al. described an individual who presented at birth with a mitochondrial disorder characterized by severe lactic acidosis and cardiomyopathy and died in the first 28 hours of life [6]. This individual was found to have a missense variant *in trans* with nonsense variant. With the limited number of cases, no true genotype-phenotype correlations can be made. However, the individuals in our study had a less severe clinical course than the individual presented by Friederich et al. [6], which may be related to the hypomorphic nature of the variant reported here that retains the ability to generate some full length *NDUFB10* transcripts. While we are confident that the deep intronic variant results in complex I deficiency, we cannot rule out that these individuals do not have a second diagnosis causing the abnormalities of the left forearm classified as a radial ray anomaly versus a secondary feature of their mitochondrial disorder as it has previously been suggested that congenital anomalies may be linked to mitochondrial dysfunction [16,17,21].

Following negative genome sequencing, RNA sequencing offers an intriguing opportunity for disease diagnosis. The family presented here had been extensively investigated previously with biochemical screening, single-gene analysis, and exome sequencing followed by genome sequencing without a molecular diagnosis being reached. Only with RNA sequencing analysis were we able to detect an overall resultant decreased expression of *NDUFB10*, and the usage of a novel splice site between exons 1 and 2 of this gene resulting from a non-coding DNA variant in *NDUFB10*. *In silico* splicing prediction tools provided only moderate confidence predictions that the identified NM_004548.3:c.131-442G>C variant would alter splicing. SpliceAI [10] gives a low confidence prediction (score=0.15) that the variant will favor the use of a donor splice site 40-nt 3’ of the variant, consistent with the position of the new donor splice site used by exon 1A. Analysis of the variant with the Human Splicing Finder tool (v3.1) [3] predicted that the variant could disrupt a hnRNP A1 Exonic Splice Silencer (ESS) motif and generate a SRp55 Exonic Splice Enhancer (ESE) motif. This highlights a specific challenge in variant interpretation, particularly in non-coding regions of the genome, where confidence is lacking in splicing predictions tools and functional annotation of the effect of genetic variants is unknown and suggests that RNA sequencing may be one approach to overcome these limitations.

In conclusion, our finding of decreased expression and aberrant splicing through RNA sequencing caused by a deep intronic variant in *NDUFB10* resulting in an isolated complex I deficiency provides further evidence of loss of NDUFB10 function as a cause of human disease. It is expected that further use of -omic testing methodologies will continue to make advances in disease diagnosis, however other methodologies warrant further study. Several recent studies have assessed the diagnostic yield of RNA sequencing in larger cohorts of affected individuals, [2, 7, 8, 12, 20] suggesting that while this approach is likely most beneficial when used in a disease-associated tissue, RNA sequencing can offer a complementary approach to existing DNA datasets and provide a further avenue for disease diagnosis. While analysis of these data and available tools are still under development, this approach may provide a new means of diagnosis in a significant number of affected individuals.

## Data Availability

The data that support the findings of this study are available on request from the corresponding author. The data are not publicly available due to privacy or ethical restrictions.

## Supplemental Data

A supplemental text file has been provided and includes a full description of methods used in this study.

## Acknowledgments

The authors thank the affected individuals and their families for their participation in this research. We acknowledge the Bio21 Mass Spectrometry and Proteomics Facility (MMSPF) for the provision of instrumentation, training, and technical support.

## Accession Numbers

ClinVar - RCV001093633.1

## Web Resources

gnomAD [11] Human Splicing Finder [3] OMIM

